# SARS-CoV-2 booster vaccine dose significantly extends humoral immune response half-life beyond the primary series

**DOI:** 10.1101/2024.02.06.24302345

**Authors:** Chapin S. Korosec, David W. Dick, Iain R. Moyles, James Watmough

## Abstract

SARS-CoV-2 lipid nanoparticle mRNA therapeutics continue to be administered as the predominant therapeutic intervention to reduce COVID-19 disease pathogenesis. Quantifying the kinetics of the secondary immune response from subsequent doses beyond the primary series, and understanding how dose-dependent immune waning kinetics vary as a function of age, sex, and various comorbidities, remains an important question. We study anti-spike IgG waning kinetics in 152 individuals who received an mRNA-based primary series and a subset of 137 individuals who then received a booster dose. We find the booster dose elicits a 71-84% increase in the median Anti-S half life over that of the primary series. We find the Anti-S half life for both primary series and booster doses drops as a function of increased year of age. However, we stress that although chronological age continues to be a good proxy for vaccine-induced humoral waning, immunosenescence is likely not the mechanism, rather, more likely the mechanism is related to the presence of noncommunicable diseases, which also accumulate with age, that affect immune regulation. We are able to independently reproduce recent observations that those with pre-existing asthma exhibit a stronger primary series humoral response to vaccination than compared to those that do not, and further find this result is sustained for the booster dose. Finally, via a single-variate KruskalWallis Test we find no difference between male and female decay kinetics, however, a multivariate approach utilizing Lasso regression for feature selection reveals a statistically significant (p-value<1×10^−3^), albeit small, bias in favour of longer-lasting humoral immunity amongst males.

## 1 Introduction

The World Health Organization has reported over 770 million SARS-CoV-2 cases and approximately 7 million SARS-CoV-2-related deaths as of December 31, 2023 (cite WHO SARS-CoV-2 dashboard). Mass vaccination remains the most effective means to prevent severity of infection and reduce the chances of hospitalization or death [1, 2]. In North America and the European Union the predominant therapeutic interventions used to reduce disease pathogenesis are the mRNA-based vaccines by Moderna and Pfizer [3]. To date, hundreds of millions of COVID-19 vaccine booster doses (defined as doses additional to the vaccine manufacturer-mandated primary series) have been administered globally [3]. Vaccine effectiveness of SARS-CoV-2 mRNA-based therapeutics wanes over time [4], thus to maintain protective immunity against severe infection, a multi-dose vaccination regiment is required [5]. It is known vaccine waning kinetics are affected by the presence of various comorbidities, age, and previous infection. Therefore, it is critical to understand how dose-dependent immunity from vaccination wanes amongst those at increased risk of severe illness from COVID-19 in order to optimize continuity of care.

Understanding dose-dependent waning kinetics is essential towards informed decision making, and targeted vaccination campaigns to higher risk populations. Although population-level efficacy studies can inform us about general trends, and are thus important towards data-informed therapeutic intervention public policy [6], the mechanism for waning occurs at the individual within-host level subject to complex immunological mechanisms [7]. Longitudinal serological studies that sample individual blood and saliva through time have been used to benchmark peak and decay responses of various SARS-CoV-2 vaccine-elicited humoral and cellular immune features [8–21]. For the mRNA-based therapeutics quantifying the adaptive immune response from multiple doses, as well as the potential comorbidic effects from noncommunicative diseases and hybrid immunity, remains an open question. To address this gap, we analyzed individual-level immune decay profiles of reported spike-specific humoral immune data for individuals who received a primary series, then subsequently a booster dose, of SARS-CoV-2 mRNA-based vaccines [21]. Our approach allows us to robustly quantify the distribution of observed decay rates following the primary series and booster doses at the individual level, thus establishing an understanding of within-host immune response heterogeneity across a cohort of individuals. We explore how decay rates following both primary series and booster dose scale as a function of age, as well as assess whether sex influences the decay profile. We further test the degree to which previous SARS-CoV-2 infection influences the decay kinetics, and further, quantify the effects of various noncommunicative diseases on the humoral decay kinetics. Overall, we find that the booster dose elicits a much longer-lasting humoral response with significantly extended humoral half lives, as compared to the primary series, regardless of which comorbidity is controlled for.

## 2 Methods

### 2.1 Data acquisition and availability

Data used for this work was acquired through a COVID-19 Immunity Task Force Grant. This data was previously published in refs. [20–22] and made available to us through the COVID-19 Immunity Task Force. Requisitions to access the data must be made through the COVID-19 Immunity Task Force.

### 2.2 Data description, model, and parameter estimation

The full data set is comprised of 152 participants. Visits 3 through 5 correspond to data collected post primary series and prior to booster dose and thus informs primary series decay kinetics. The data in this time frame has 151 individuals comprising a total of 425 data points. Booster dose decay kinetics are captured on visits 6 through 8 in a subset of 137 individuals, comprising a total of 399 data points. This time frame corresponds to data collected post booster dose, prior to a fourth dose.

Mathematical models have coupled various individual-level immune features, such as cytokines, plasma B cells, and IgG/IgM, into a coherent mechanistic modelling structure in order to model vaccine durability [23–25]. Such modelling approaches have also been used to determine which immunological features drive the timescales of vaccine uptake, immune priming, peak humoral immunity, as well as humoral decay, thus elucidating key variables inherent to vaccine-elicited immune dynamics [26]. Modelling has also been used to determine how vaccine-driven IgG counts decay as a function of age, prior infection, T-cell response, and dosing intervals [27]. Here we employ a simple exponential approach to assess immune waning decay kinetics. The model used in this work is

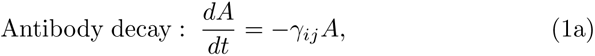

where *A* represents IgG values, and *γ_i,j_* is the decay rate for the *i*^th^ person for the *j*^th^ dose regimen. Here, *j* is either 2, corresponding to the primary series decay rate for the *i*^th^ person, or 3, corresponding to the booster dose decay rate for the *i*^th^ person. From Eq.1 the half lives,*τ_ij_*, of each individual for the primary and booster doses are derived as

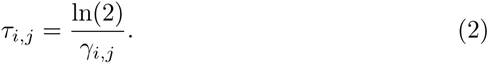

All model fits were performed in Monolix [28] (Version 2020R1) using non-linear mixed-effects models. Individual parameters for each data set are determined by the maximum likelihood estimator Stochastic Approximation Expectation–Maximization (SAEM), and all fits met the standard convergence criteria (complete likelihood estimator). Using this modelling approach allows us to leverage the statistics of the underlying data set to optimally learn predicted trends and fitted variables. All individuals are simultaneously fit, where key differences between Anti-S responses are determined through leveraging various statistical models utilizing both fixed and random effects. We report relative standard error of the parameter values, and further report relative standard error of the random effects which assess practical identifiability. We employ two separate fits in Monolix: one fit across all primary decay kinetics, thus resulting in 151 individual fits as well as a single population fit, and a second fit across all booster decay kinetics, thus resulting in 137 individual fits as well as a single population fit. For each of the fits we also fit for the initial condition in Anti-S which is typically determined to be near-equivalent to the initial value being fit to. Further details regarding the observation and error models used in this work can be found in supplementary section S2. Fig. 1A displays an example trajectory for Anti-S as a function of time for a single individual who received three doses of BNT162b2. Dashed lines indicate vaccine inoculation times.

**Fig. 1.**
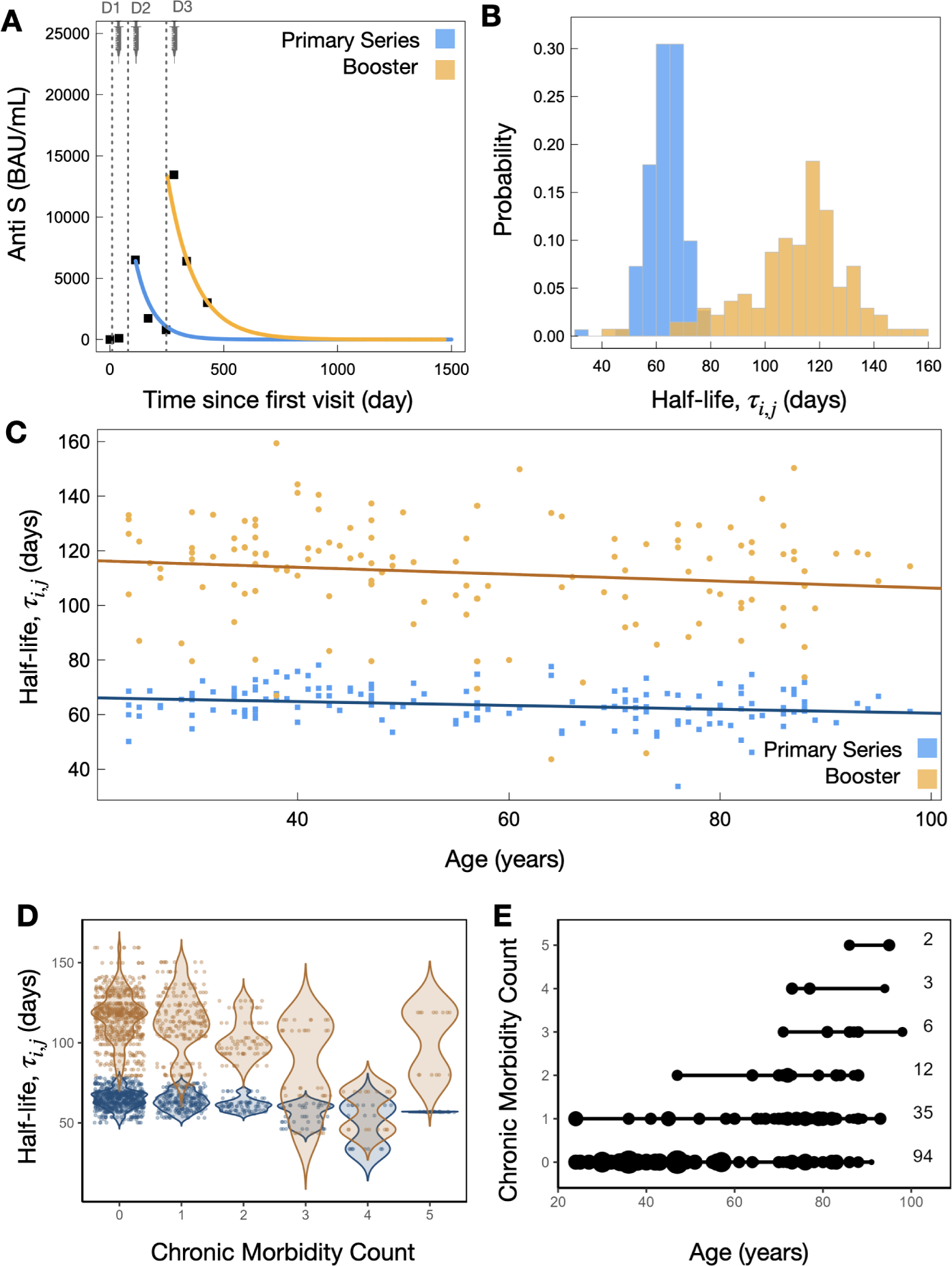
A) Example individual model fits (Eq. 1) for the primary series (blue) and booster dose (orange) Anti-S trajectory data. B) Distribution of all individual half lives (Eq. 2) for primary series (blue) and booster dose (orange). C) Individual Anti-S half life as a function of chronological age for primary series and booster dose, where a decreasing trend as a function of increased age is found. D) Violin plots of Anti-S half lives and number of chronic morbidities. Points are plotted with a jitter function to display the density of individuals with each chronic morbidity count. E) Plot of chronic morbidity count and chronological age. The size of the point represents the number of individuals at a particular age who share the same chronic morbidity count. Where the smallest points represent one individual and the largest points represent seven individuals. The counts for each chronic morbidity category are provided to the right of the figure.

### 2.3 Statistical analysis

We employ the Kruskal-Wallis Test [29] to compute cross-sectional comparisons between two groups within our categorizations of sex, age and number of doses. The pairwise Wilcox test is used to make multiple group comparisons between healthcare workers, residents and seniors. To compare p-values they are adjusted with the Benjamini-Hochberg method [30]. Statistical tests are carried out in *R*, version 4.2.2 (2022-10-31). On our violin plots, we indicate the p-values between cross-sectional group comparisons by using the following legend: ‘*’ for 0.01 *< p ≤* 0.05, ‘**’ for 0.001 *< p ≤* 0.01, ‘***’ for 0.0001 *< p ≤* 0.001, and ‘****’ for *p ≤* 0.0001. Multivariate linear regression (MVLR) was employed to elucidate the interplay between age and vaccine-induced immune response. The objective was to quantify the influence of age on the variability of antibody decay rates, controlling for confounding variables such as chronic comorbidities and other demographic factors. This methodological approach allows for a nuanced analysis of the factors that contribute to individual variations in antibody decay rates.

#### 2.3.1 Variables and Factor selection

This section outlines the specific methodologies, variables, and criteria employed in our multivariate linear regression analysis. The variables considered can be found in Table 1.

**Table 1.** Variables included in the analysis for both primary series and booster dose. Dummy Variables for Comorbidities and Natural Infection: Presence (1) indicates the Reference Category; Absence (0) denotes the Baseline Variable.

| Variable | Description |
| --- | --- |
| Anti-N Result | SARS-CoV-2 Nucleocapsid |
| Anti-Spike Censored | binary variable for upper limit of quantification for Anti-S |
| Type_short | Short type code (occupation/category) |
| Sex_at_Birth | Sex of the participants at birth |
| ageMinMaxNormalized | Chronological age of participants min-max normalized |
| Chronic Neurological Disorder | Presence of chronic neurological disorders |
| Hypertension | Presence of hypertension |
| Asthma | Presence of asthma |
| Chronic Lung Disease | Presence of chronic lung diseases |
| Cancer | Presence of cancer |
| Predicted Infection (Primary Series) | A constructed variable indicating the presence of natural infection in the primary series time-frame |
| Predicted Infection (Pre Booster) | A constructed variable indicating the presence of natural infection |

Our MVLR model was developed in R version 4.2.2 (2022-10-31 ucrt). For feature selection, Lasso regression was employed using the glmnet 4.1-7 package [31–34]. Cross-validation was performed with cv.glmnet, incorporating standardization to ensure the model’s robustness and the optimum *λ* was used to decide feature inclusion. Lasso penalization, a L1 regularization technique, incorporates a penalty term equal to the absolute value of the magnitude of coefficients into the loss function minimized during regression.As *λ* increases, more coefficients are set to zero leading to feature selection. For more detail see the supplementary section S5.1.

The variable ‘Anti-Spike-Censored’ was created to identify instances where the anti-SARS-CoV-2 RBD antibody titers exceeded the upper limit of quantification (ULOQ) after log transformation of the Enzyme-Linked Immunosorbent Assay (ELISA) data. This binary indicator was incorporated into our lasso and multivariate regression analyses to adjust for censored observations in the dataset.

The correlation between factors was calculated for the chronic conditions, demographic factors, and vaccine dose responses (Fig.S7, Table.S1).The heatmap in Fig. S7 uses color intensity to represent correlation strength: red for positive, blue for negative, and white for no correlation. A Variance Inflation Factor (VIF) analysis was also conducted to assess multicollinearity among the regression model variables (Fig.S8). Except for ‘ageMinMaxNormalized’, all variables presented VIF values below the conventional threshold of 2, indicating minimal multicollinearity concerns [35–37]. Despite a marginally high VIF value, ‘ageMinMaxNormalized’ was retained in the model due to its analytical significance.

Fig. S5 presents the distribution of chronic conditions within our study population, depicted as the percentage of total diagnoses for each condition. Hypertension was the most prevalent comorbidity, accounting for 35 cases, followed by diabetes with 17 cases. Chronic heart disease and chronic neurological disorder were also notable, with 14 and 13 cases respectively. Asthma was identified in 11 individuals, while both chronic kidney disease and chronic lung disease were present in 7 participants each. There were 6 cases of cancer and 2 cases of chronic blood disorder.

To asses the potential confounding effects stemming from comorbidity overlap within our cohort, hierarchical clustering was applied. We use the ‘hclust’ function in R, which uses the “complete” linkage method. In this approach, the distance between two clusters is defined as the maximum distance between any two points in the clusters. The resulting patterns are depicted in Fig. S6. Additionally, to quantify the likelihood of concurrent chronic conditions, we computed conditional probabilities, as illustrated in Fig. S9. The clustered heatmap in Fig. S6 reveals the co-occurrence of chronic conditions, with blue cells representing the presence of comorbidities and the dendrogram indicating clustering based on their co-occurrence frequency. These analyses enabled the detection of distinct clusters of comorbidities, which may affect the estimated decay rates of vaccine-induced immunity. Hierarchical clustering allowed us to systematically evaluate the interplay of multiple chronic conditions, which is critical in ensuring the robustness of our findings against the confounding effects of comorbidity overlap.

The identification of participants with prior natural SARS-CoV-2 infection is complicated by antibody waning. Over time, the concentration of antibodies can diminish, leading to a decrease in detectable levels, particularly for the anti-N antibodies associated with natural infections. This waning complicates the distinction between vaccine-induced immunity and hybrid immunity – a combination of natural and vaccine-induced immune responses. To assess the likelihood of prior COVID-19 infection, a variable named ‘predicted infection’ was created based on the quantified levels of N-terminal domain (anti-N) antibodies. Given that anti-N antibody levels are indicative of a natural infection rather than a vaccine-induced response, we established thresholds to categorize the likelihood of prior COVID-19 infection (Fig. S4).

- **High Threshold:** This threshold was set based on the maximum anti-N antibody level found in participants with no recorded positive COVID-19 test, thus establishing the upper limit of antibody levels in individuals without a history of infection.
- **Low Threshold:** Specifically chosen for the ‘predicted infection’ variable, it was calculated as twice the lowest anti-N antibody level observed in individuals with confirmed COVID-19 positivity, intended to capture those with lower, yet detectable, antibody levels suggestive of past infection.

Participants with antibody levels surpassing the established Low Threshold were considered to have potential hybrid immunity, reflecting the likelihood of a previous natural infection combined with vaccination. Our methodology acknowledges the challenge of accurately discerning natural infection solely based on serological data due to the temporal decline in antibody levels post-infection. In the analysis, rather than filtering out potential cases of hybrid immunity, we retain all data and introduce the ‘predicted infection’ variable. This is done to retain data and to examine the influence of hybrid immunity on the estimated decay rates of vaccine-induced immunity.

## 3 Results

### 3.1 Booster doses significantly extend humoral half life as compared to the primary series

In Fig. 1A model-fit predictions are extended out to illustrate model-driven long-term projections. Additional individual fits are displayed in Fig.S1, with the cohort-level population fit illustrated in Fig.S2. Fig. 1B displays the distribution of half lives determined from Eq.2 for both the primary series and booster doses. We find the distribution of half lives between primary series and booster dose to have little overlap, with clearly differing median and mean responses. For the primary series fits across the 151 individuals fit in this work we find a median Anti-S half life of 63.3 days with interquartile range (IQR) 7.9 days. For the booster dose fits across 137 individuals we find a median Anti-S half life of 115 days with IQR 20 days. Therefore, taking the IQR range into account, the booster dose leads to a 71-84% increase in Anti-S half life over that of the primary series.

### 3.2 Vaccine immune waning as a function of age and chronic morbidity count

We find a linearly decreasing trend in Anti-S half-lives with increased age for both the primary series and booster dose (Fig. 1c). For both the primary series and booster dose in Fig 1c we plot the Anti-S decay half lives as a function of chronological age where we find a linearly decreasing trend in Anti-S half life as a function of increased age. A linear fit of the individual decay rates with age reveals that half life decreases by 0.1 days (or 2.4 hours) per year of age for the primary series (blue) and 0.13 days (or 3.12 hours) per year of age following the booster dose (orange). In a separate analysis in Fig. 1d, we plot the individual decay rates as a function of the number of chronic comorbidities reported by the individuals in the study. We find that the Anti-S decay rate increases with zero through to four self-reported comorbidities, while for 5 comorbidities the decay rate is observed to decrease relative to those with four self-reported comorbidities. In Fig. 1e we present a parametric plot of chronic morbidity count and individual age. Here, the size of the point represents the number of individuals at a particular age who share the same chronic comorbidity count. Where the smallest points represent one individual and the largest points represent seven individuals. This parametric figure demonstrates the distribution of chronic comorbidities in relation to age within the study’s cohort, with the majority of individuals exhibiting 0 or 1 comorbidity, and also displays a modest positive correlation between an increasing age and chronic comorbidity count.

In a separate analysis in Fig. 1d, we plot the individual decay rates as a function of the number of chronic comorbities reported by the individuals in the study. We find that generally the Anti-S decay rate increases with increasing numbers of reported comorbidities. Fig. 1e further explores the relationship between comorbitities and age by depicting a parametric plot of chronic morbidity count against individual age. The plot uses varying point sizes to indicate the number of individuals at each age with similar chronic morbidity counts, ranging from one to seven individuals per point size. Fig. 1cde suggest a relationship between chronic comorbidity and age that warrant further exploration: Panel c suggests a link to age, Panel d considers the potential primary influence of comorbidity, and finally, Panel e highlights the relationship between comorbidity and age.

### 3.3 Statistical comparisons between sex, age, and status (HCW, senior, resident)

Single-variate statistical comparisons between the primary series and booster dose Anti-S decay rates across the entire population reveal significant p-values of 2.2×10^−16^ (Fig. 2A, left panel, blue and orange colours), demonstrating there is a clear difference in decay profile between primary series and booster dose. In contrast, we find no significant differences when stratifying for sex within primary series or booster dose decay rates when assessing by a single-variate test (Fig. 2A, middle panels, purple and and green colours).

**Fig. 2.**
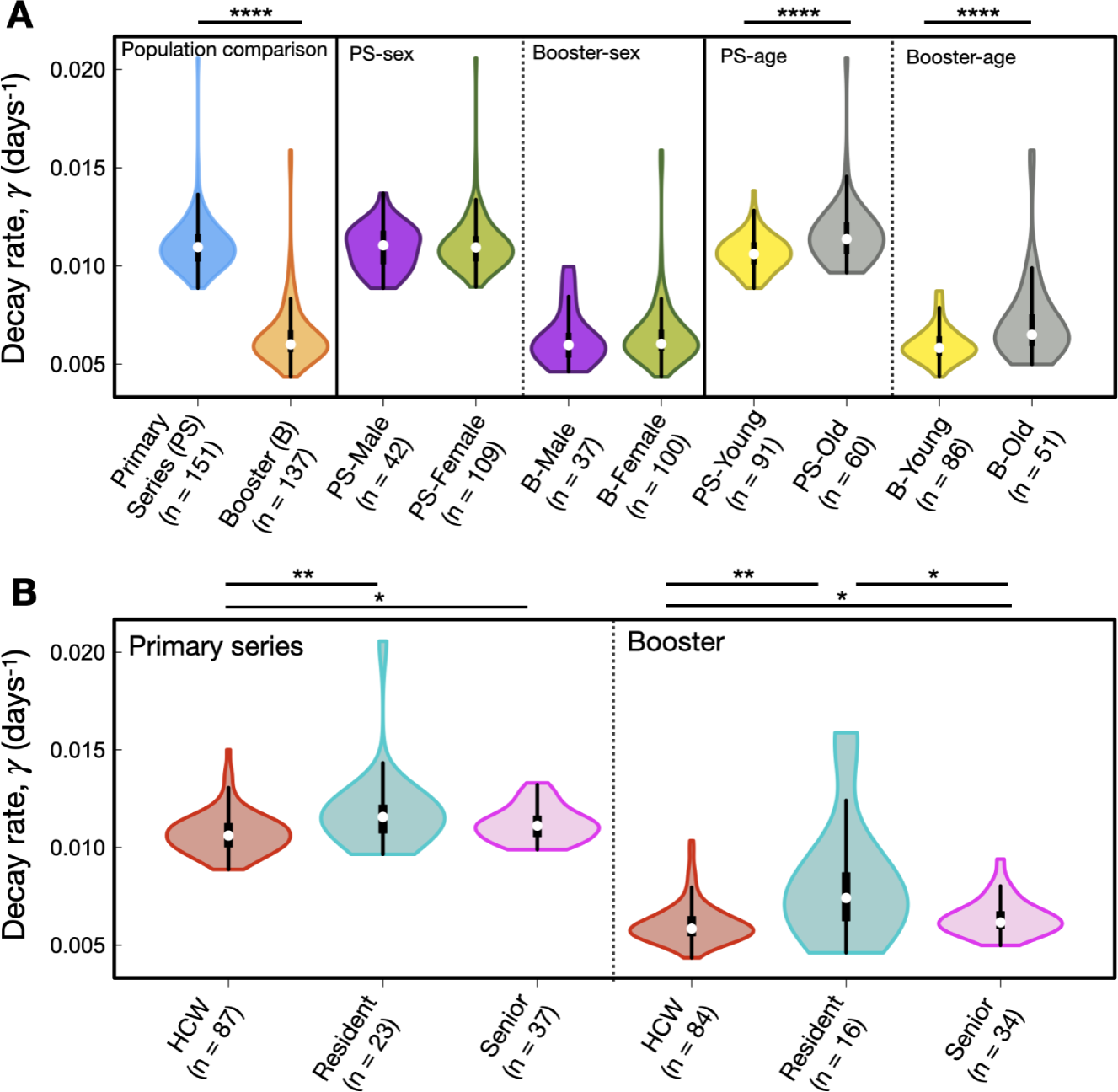
A) Cross-sectional statistical comparisons. In order from left to right: population comparisons between primary series and booster dose decay, primary series sex-based comparison, booster sex-based comparison, primary series age-based comparison, booster age-based comparison. B) Left: statistical comparisons between HCW, residents and seniors for primary series decay rates. Right: statistical comparisons between HCW, residents and seniors for booster-elicited decay rates. We note the y-axis is decay rate, which is inversely proportional to half life (see Eq. 2)

The Kruskal-Wallis test between young and old individuals requires a prescribed boundary between young and old. To test for the influence of this age boundary on age-stratified distributions of decay rates we varied the age boundary from 40 to 75 years of age. Those people with ages below the boundary are considered ‘young’ while those above the boundary are considered ‘old’. For age boundaries of 40-75 years of age Fig. S3 displays all Bonferoni-corrected p-values. We find that for the primary series there is a number of age boundaries of 58-70 years of age that lead to roughly equivalent minimal p-values less then 1×10^−3^. Similarly, For the booster dose we find that young/old age boundaries from 52-70 years of age lead to a number of minimal p-values in the range of 10^−3^-10^−4^ (See Fig. S3). For illustrative purposes, we provide one such comparison in the right-hand panel of Fig. 2A shown in yellow (young) and grey (old) colours for an age cutoff at 70 years, noting that age cutoffs of between 52 and 70 years of age lead to roughly equivalent p-values (Fig S3).

In Fig. 2B we explore comparisons between health care workers (HCWs), residents, and seniors, with median ages of 41, 86, and 77, respectively. Where HCW tend to be significantly younger than residents and seniors it is therefore unsurprising that we find significant differences between both HCW and residents and seniors for both the primary and booster series. However, it is not necessarily surprising that HCW display significantly different decay kinetics as compared to residents and seniors. SARS-CoV-2 poses an occupational health risk to HCWs who may be repeatedly exposed to the virus [38]. Therefore, in the next section we consider a multivariate regression analysis that simultaneously considers variables such as age, occupation, Anti-N levels, comorbidity count, and sex, to see which variables are driving Anti-S decay kinetics.

### 3.4 Multivariate analysis to determine factors driving decay kinetics

In previous sections, we reported model-determined individual antibody decay rates and half-lives, demonstrating their correlation with age and cumulative comorbidity count. Both factors have proven to be reliable proxies for predicting trends in Anti-Spike (Anti-S) antibody decay kinetics.

Building upon these findings, we have conducted a MVLR analysis to parse out the individual effects of the comorbidities that constitute the overall comorbidity count. Fig. 1e illustrates a notable correlation between age and comorbidity count. The application of MVLR enables us to explore the interplay between age and chronic comorbidities, elucidating the impact of immunosenescence—the age-related decline in immune function—after adjusting for coexisting chronic conditions. This analysis allows for more nuanced understanding of the factors effecting Anti-Spike (Anti-S) antibody decay kinetics.

The MVLR analysis revealed a complex landscape where various covariates exert differential impacts on the decay kinetics of Anti-S antibodies, Fig 4. Notably the model highlighted both the biological and statistical significance of chronic lung disease, showing an increased antibody decay rate with both the primary and booster series.

For the booster dose, the model highlighted chronic lung disease as a significant predictor of antibody decay rate, with an estimated regression coefficient of 1.873×10^−3^ (with an estimated p-value of less then 1×10^−6^). This was consistent with the primary series results, where chronic lung disease also showed a notable positive association with antibody decay rate, albeit with a slightly lesser effect size, with an estimate of regression coefficient of 4.135×10^−3^ (p-value less then 1×10^−6^). The presence of hypertension (estimated coefficient of 0.325×10^−3^, p-value = 1.701×10^−2^) also indicated an increased decay rate in the booster dose analysis. However, for the primary series, while a positive trend was observed, the results did not reach statistical significance.

Conversely, asthma was associated with a slower antibody decay rate in the booster dose (estimated coefficient = *−*0.783×10^−3^, p-value = 1.5×10^−5^). Interestingly, the age variable, indicated as ageMinMaxNormalized, showed a positive effect on the decay rate for the booster dose (estimated coefficient 0.846×10^−3^, p-value = 1.777×10^−2^), suggesting that older age may be associated with a more rapid decline in antibody levels post-booster.

The model’s intercept for both doses indicated significant baseline decay rates (Booster Dose: estimated coefficient 6.06×10^−3^, p-value less then 1×10^−6^); Primary series: estimate 1.801×10^−3^, (p-value less then 1×10^−6^)), suggesting inherent decay that occurs independent of the studied covariates.

Sex at birth, presented as a statistically significant factor in both the booster dose (estimated coefficient 0.367×10^−3^, p-value = 7.13×10^−4^) and primary series (estimated coefficient = 0.214×10^−3^, p-value = 2.44×10^−2^), indicating a sex-based difference in antibody longevity. This finding are further elaborated in the discussion section.

Within our dataset, the Anti-N variable produced near-zero estimates for both the primary series (estimate coefficient = 0.005×10^−3^, p-value = 8.629×10^−2^) and the booster dose (estimate coefficient = 0.007×10^−3^, p-value = 3.457×10^−2^), accompanied by an estimated standard error of 3×10^−6^ for both primary series and booster dose. This suggests an insignificant direct impact of Anti-N on antibody decay rates with the predicted infection variable included.

In our analysis, we introduce two key variables: ‘predicted infection (pre booster)’ for the primary series and ‘predicted infection’ (any phase) for the booster dose, see Sec. 2.3.1 for details. These variables are incorporated into the regression model to assess the effects of potential hybrid immunity. This approach allows for the consideration of hybrid immunity, taking into account the possible influence of natural infections during the periods of estimated antibody decay rates. We see a significant influence of hybrid immunity in the data (estimate coefficient = 0.419×10^−3^, p-value = 3.591×10^−3^) with the primary series and (estimate coefficient = 0.583×10^−3^, p-value = 9.65×10^−4^) with the booster dose.

In the analysis of the primary series, being a resident and being a senior emerged as significant factors, with positive associations to antibody decay rates (resident estimate coefficient = 0.915×10^−3^, p-value *<* 1×10^−6^; senior estimate coefficient = 0.381×10^−3^, p-value = 2.06×10^−4^). However, in the booster series, we observed a different pattern, where the age variable (ageMinMaxNormalized) is retained by the lasso penalization with a positive association on the estimated decay rate (estimate coefficient = 0.846×10^−3^, p-value = 1.777×10^−2^).

To explore changes in the importance of age and type factors, we consider the lasso regression analysis. Considering the lasso regression analysis in Fig. 3 the booster series analysis, the coefficients for resident and senior status diminish at a smaller penalization value in the lasso path, while the age variable the age variable continues to be non-zero past a lambda penalization value 1 standard deviation above the optimal *λ*. This suggests a shift in the relative importance of these variables when considering the primary as compared to the lasso analysis considering the booster dose. When considering the lasso analysis of the primary series we observe an opposite trend where the age variable’s influence diminishes as the lasso penalty increases, leading to a pronounced change in coefficients for resident and senior status.

**Fig. 3.**
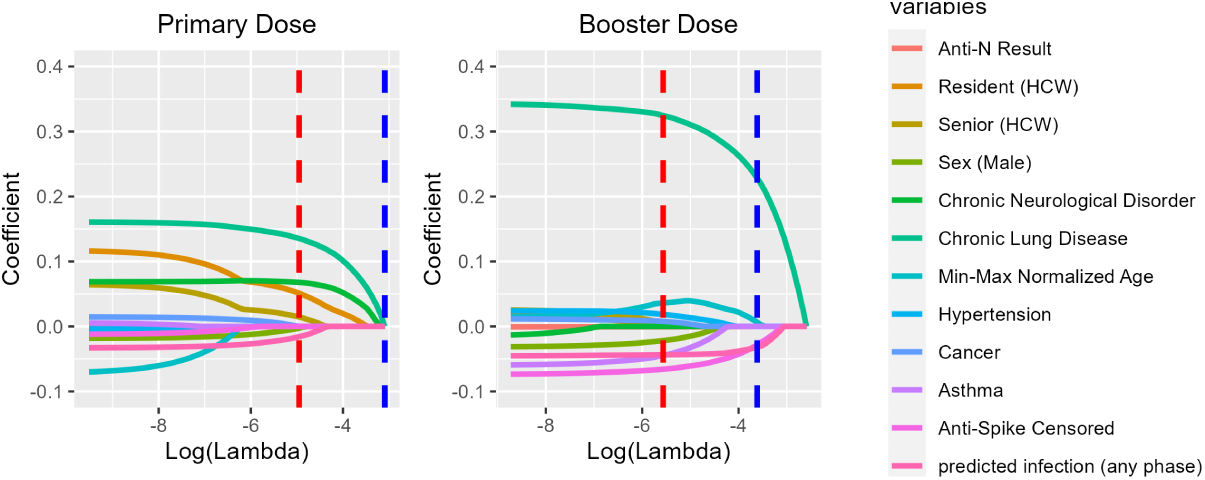
Lasso regression coefficients for Primary and Booster Dose responses across various health and demographic variables. Each color-coded curve corresponds to a specific variable, illustrating its coefficient value as the regularization parameter (*λ*) changes. The red dashed line represents the optimal *λ* (min *λ*) where the model demonstrates the best fit, while the blue dashed line denotes a *λ* that is 1 standard deviation away from the minimum.

The observed strong positive correlation between age and senior status (correlation coefficient = 0.62) implies that the effects of age and senior status are not distinct. The influence of age seems to partially subsume or displace the effects associated with being a senior or a resident, especially when examining the booster dose data. This analysis, conducted using the lasso method for feature selection, underscores the limitations of our current dataset and the changing population dynamics between the two groups. Given a consistent sample, we would expect a consistent pattern in the influence of age across both series - either diminishing or remaining significant in both. However, due to these constraints, we observe divergent results in the impact of age, residency status, senior status, and comorbidities on the decay rate of antibodies, highlighting the complexity and interdependence of these factors.

Our multivariate linear regression analysis has unveiled a complex immune landscape where factors such as chronic lung disease, hypertension, asthma, along with demographic variables like sex, age, and residency status, significantly influence Anti-S antibody decay rates. These findings highlight the intricate interplay between individual health conditions and demographic characteristics in shaping the body’s response to vaccination.

## 4 Discussion

We find a narrow distribution of individual Anti-S half lives following primary series, with median value of 63(IQR:7.9) days (Fig. 1b). Following the booster dose, the median half life is significantly increased, as well as the dispersion of half lives around the median. We find a booster dose median half life of 115(IQR:20) days (Fig. 1b). Thus, our study is in agreement with previous work [19, 39] that SARS-CoV-2 booster doses lead to significantly longer-lasting humoral immunity as compared to the primary series. Where this current study is predominantly concerned with the BNT162b2 vaccine, it is interesting to note that these booster-extended half lives are found in a cohort who received a mixture of vaccines [19] as well as those who received inactivated virus vaccines [39], suggesting booster-extended humoral half lives are a universal feature of a multi-dose SARS-CoV-2 vaccination regimen. Matveev *et al.* [19] followed a cohort of individuals for over 300 days post dose one, who then received three SARS-CoV-2 vaccination doses, and found spike-specific memory B cells monotonically increased over the study timeline [19]. Thus, it may be that the timescales of SARS-CoV-2 vaccine inoculation are such that persistent new antigen-specific PCs are recruited while previous antigen-specific PCs have yet to diminish leading to humoral immunity to be bolstered by increasing amounts. This hypothesis is further supported by recent findings on bulk plasma B cell dynamics in mice inocculated with a model antigen which suggest a long-lived plasma cells recruited through the germinal centre have a half life 700 days while NP-specific persistent plasma cells display a half life of 23 days [40]. Another mechanism potentially contributing to increased antibody half life following subsequent vaccinations may be related the recent discovery that SARS-CoV-2 mRNA vaccine sequences have been found to circulate in the blood up to 28 days post vaccination [41]; a significant increase over previous estimates of 48hrs (human breast milk) [42] and 15 days (qPCR of human blood samples) [43]. Thus, immune system priming may occur over extended periods of time following each inoculation, and may not have ceased before subsequent doses are administered.

Anti-S and Anti-RBD immunogenic outcomes following the primary series and booster doses from mRNA-based SARS-CoV-2 vaccines have been previously identified in humans. In a study of 3407 individuals who received the BNT162b2 primary series, Aldridge *et al.* [44] found an anti-S half life of 72 days, and found no evidence of age or sex-related trends. Similarly, in a study of 3259 older individuals in long-term care no differences between male and females in anti-S-IgG titers over time were found, albeit, slightly more local vaccine-induced adverse effects were reported in females [45]. In an aged 50+ cohort of individuals, Matveev *et al.* [19] find IgG spike half-life values of 28.9*±*7.2 days following the primary series, with the half-life increasing substantially post booster dose to 80*±*22.4 days. A study on young (age 31-40) individuals who received three inoculations of the inactivated virus CoronaVac vaccine found antibody half lives of 28(95% CI: 26-32) days following the primary series, which increased three-fold to a half life of 83(95% CI:64-119) days following the booster dose [39].

From the literature, spike IgG from infection is typically found to have a significantly longer half life as compared to that following two doses of vaccination. Spike IgG half lives of 126 days [18], 110 days (amongst males [9]), and 159 days (amongst females [9]) have been reported. Mild, moderate, and severe COVID-19 infection elicits differing immunogenic responses and may be correlated with comorbidities and influenced by sex [46–48]. Thus, the influence of hybrid immunity from vaccination and infection is an important variable. Anti-N levels are often utilized to assess for COVID-19 infection as they are not generated by vaccination [**?**]. Here, we use patient Anti-N levels in our multivariate statistical tests to classify individuals as infected or not (see methods for details). Consistent with previous literature, individuals classified as infected by their Anti-N values were determined to have significantly slower Anti-S half lives, and thus a longer-lasting humoral response (Fig. 4). Accordingly, we included remaining individuals who were not classified as infected and included their Anti-N levels in our multivariate analysis, whereby the Anti-N levels of these individuals classified as “uninfected” have no statistical influence on Anti-S half lives.

**Fig. 4.**
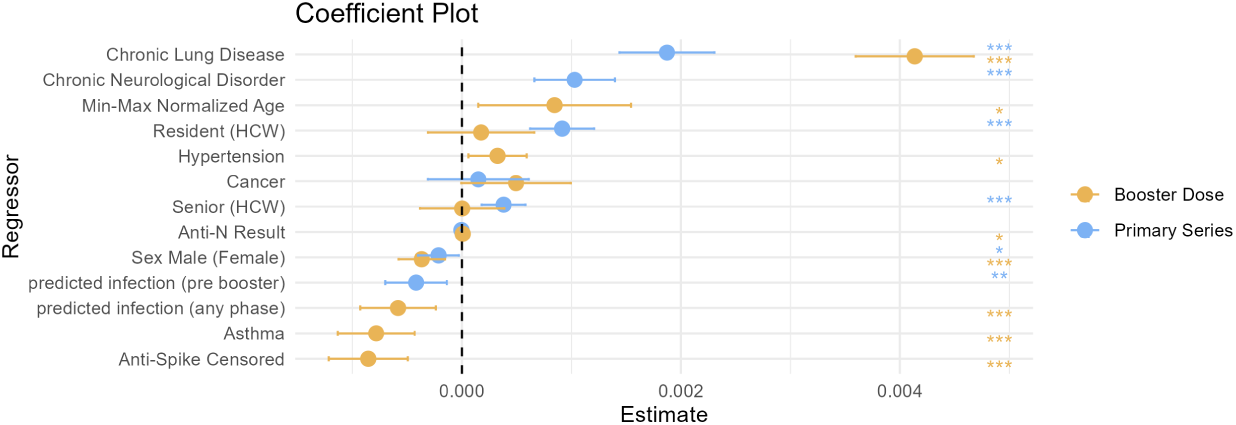
The outcomes of the multivariate regression analysis, conducted with factors selected via lasso regression at the optimal lambda value, are illustrated in Fig. 3. This figure includes 95% confidence intervals for each coefficient. The primary and booster series are depicted in blue and gold, respectively. Statistical significance is indicated by stars: ‘***’ for *p <* 0.001, ‘**’ for *p <* 0.01, and ‘*’ for *p <* 0.05. On the x-axis, the effect size is centered around zero; positive values to the right suggest an increased antibody decay rate compared to the reference category, while negative values to the left suggest a decrease. The magnitude of the associated effect is proportional to the distance from the center line. The reference category is specified in brackets. For dummy variables, each coefficient represents the change in the outcome relative to this reference within the categorical factor. For a table of numerical results see Tab. S1.

In our single-variate statistical estimates employing the Kruskal-Wallis Test on the distribution of decay rates, we find no differences between males and females for both booster and primary series (Fig. 2A). However, our multivariate regression analysis, which simultaneously compares all variables of interest on their influence over decay kinetics, reveals sex at birth to be a significant factor for antibody longevity for both the primary series and booster doses, with males displaying slightly slower Anti-S decay kinetics as compared to females (Fig. 4). We stress that although the multivariate p-value is less than 10^−3^, the males display only a slightly slower decay response than compared to females. Thus, our multivariate test reveals males to have only slightly slower decay than compared to females. Genetic and biological mechanisms leading to differences in vaccine-induced humoral immunity between sexes have been previously discussed in the literature [49], with SARS-CoV-2 vaccination examples noted in the previous paragraph.

We find our fitted decay-rates show several differences from previously reported studies using iterations of this same data set [20–22]. Brockman *et al.* [21] originally report a half life of IgG RBD of 87(95% CI: 75-97) at 3 months post primary series of vaccination in the COVID-19 naive group. In a follow up paper utilizing an additional point at 6 months post primary series, as well as filtering the data by “restricting the analysis to participants with a complete longitudinal data series with no values above the ULOQ”, the Anti-S half life were found to drop to a median value of 59(IQR: 52-75) days amongst HCW and 52(IQR:45-65) days amongst older adults, thus validating that humoral immunity is declining faster in older individuals [20]; these values are very close to our population median estimate of 63(IQR:7.9) days. In a follow up study on the same individuals [22], estimates of WT-specific Anti-RBD IgG half lives amongst COVID-19 naive individuals post booster dose were found to be 73(IQR:53-101) days in HCW and 69(IQR:54-91) days amongst older individuals. For Omicron-BA.1-specific IgG they find a half life of 75(IQR:58-93) days in HCWs and 78(IQR:64-94) days amongst older individuals, whereas we find a population median booster-elicited half life of 115(IQR:20) days [22]. In our work, we employ non-linear mixed effect models to fit Anti-S humoral data. We do not filter for individuals who have data points on the ULOQ, or suspected hybrid immunity at the stage of fitting. Rather, we fit all the humoral decay data points on all individuals, as well as on the entire population, and employ a multivariate linear regression analysis on the fit-determined decay rates to examine which factors have statistical explanatory power in driving decay rates to be faster or slower. We further include in our test whether the data points on the ULOQ are significantly effecting the observed decay kinetics. Previous studies reported that individuals with data points on the ULOQ lead to an underestimate of the decay rate [19]. Consistent with previous work, shown in Fig 4 we find that Anti-S ULOQ censored data points do indeed lead to an underestimate of decay-rates, which we attempt to consider and correct for in our multivariate statistical approach. Differences between our fitted decay rates, versus those previously fit with this data set, may be due to data filtering and the algorithm employed to fit the data. The details of our fitting algorithm can be found in the methods section and supplementary sections S1 and S2.

We find the variable with strongest explanatory power leading to rapid humoral decay, following both the primary series and booster dose, is the presence of chronic lung disease, while the strongest variable leading to sustained humoral immunity is pre-existing asthma. Previous reports following a cohort of individuals with chronic pulmonary diseases who received a primary series of BNT162b2 have found impaired humoral immunological outcomes as compared to a control [50]. In another study, Li *et al.* [51] follow 1400 patients with chronic disease, as well as 245 healthy controls, who received either a SARS-CoV-2 protein subunit recombinant vaccine or inactivated virus vaccine, and find that those with chronic lung disease display amongst the lowest median RBD-IgG and NAbs titres as well as the lowest maximum range in antibody response as compared to the other diseases. Furthermore, Liu *et al.* [52] find that SARS-CoV-2 vaccine-elicited immunity amongst those with chronic lung disease leads to significantly reduced SARS-CoV-2 vaccine antibody titres and decreased vaccine-specific memory B-cells. It is therefore unsurprising that we find chronic lung disease correlates with rapid a humoral decay kinetics; however, it is surprising to find that chronic lung disease leads to (statistically) significantly faster Anti-S decay than age, hypertension, cancer, and chronic neurological disorder. Where chronic cigarette smoking has been found to reduce antibody titres to vaccination, and lead to more rapid vaccine-elicited humoral decay [53], we note that only a single individual (ID 122) with chronic lung disease reported as ‘yes’ to being a smoker of cigarettes. Thus, potential affects of chronic smoking on dysregulation of adaptive and innate immune function [54] cannot explain our result.

For those with existing asthma, respiratory infections are more likely to be severe [55], which means it is generally important for these individuals to get vaccinated against respiratory diseases. The relationship between SARS-CoV-2 infection severity and the presence of immune-mediated inflammatory diseases, such as asthma, is complex. For example, initial SARS-CoV-2 reports revealed, rather unintuitively, a low prevalence of asthma among patients with severe COVID-19 [56]. Allergic diseases, such as asthma, are characterized by increased levels of T-helper cell type-2 (Th2) which secrete cytokines associated with airway inflammation [**?**]. A recent SARS-CoV-2 LNP mRNA-based study has demonstrated that people with asthma who are on cytokine inhibition drugs respond more poorly, compared to a healthy asthma-free control group, to SARS-CoV-2 vaccination [57]. Humoral immunity was found to be significantly lower amongst individuals with immune-mediated inflammatory diseases on an anti-TNF drug regimen compared to a healthy control, while those with an immune-mediated inflammatory diseases that was untreated had significantly higher RBD and spike responses than compared to the healthy control [58]. Further, people with allergic rhinitis (AR) were found to have a significantly stronger humoral response to SARS-CoV-2 vaccination than compared to otherwise healthy individuals; this outcome was hypothesized to be due to the enhanced type 2 follicular helper T (TFH2) cells in those individuals with AR [59]. It is known people with asthma, in the absence of cytokine inhibiting therpapies, have elevated TFH2 concentrations [60, 61]. TFH cells provide help to B cells which then cascades into the humoral response [62], it is therefore suspected that the elevated levels of TFH cells present in individuals with untreated immune-mediated inflammatory diseases, leads to elevated B-cell priming and therefore a more robust humoral response. Cytokine-inhibiting therapies often used to treat immune-mediated inflammatory diseases may inhibit the TFH-B-cell interaction and therefore lead to a less robust humoral response [58]. Our study contains 11 individuals with asthma, where none of the individuals reported being on immune-suppressant drugs. Consistent with previous literature, our multivariate statistical analysis reveals that the individuals with asthma display significantly slower humoral decay (Fig. 4). We suspect this result is due to elevated TFH responses due to the presence of asthma, however, these cell types were not assessed in our data set.

It is clear from previous studies that age plays a critical role in humoral immunity elicited from SARS-CoV-2 mRNA-based vaccines [8, 23, 63–65]. The faster decay rates observed as a function of increased age, in the absence of previous infection, may be explained by the natural process of immunosenescence, defined as the collective diminishment of humoral and cellular immune responses as a function of age [66]. Indeed, previous studies have discussed implications of immunosenescence in mounting immune responses from the predominant SARS-CoV-2 vaccines [67, 68]. We find primary and booster dose Anti-S half-lives steadily decrease with increased age (Fig. 1c). Further, our results show that a simple single-variate statistical test reveal significant differences in decay kinetics when grouping by ‘young’ and ‘old’ (Fig. 2a and Fig.S3). Immunogenicity from vaccination in older patients is complicated by those who are hybrid immune or breakthrough infectious; no correlation with spike-specific antibody and age has been found in these cases from previous studies [63]. Further, our ‘young’ cohort is predominantly composed of HCWs. HCW’s Anti-N and Anti-S decay profiles may be distinct from the general population as HCWs may be regularly exposed to SARS-CoV-2 [69, 70]. The ‘old’ cohort in this study is composed of residents who reside within longterm care or assisted living facilities, and ‘seniors’ who live independently. There are known to be complex heterogeneous SARS-CoV-2 disease burdens amongst older populations [71]. Accumulated noncommunicable disease with increased age may interfere with SARS-CoV-2 immune responses [72]; where various noncommunicable diseases may influence vaccine-induced immune profiles [50–52, 73]. The single-variate estimate in Fig. 2a is “näıve” in the sense that diseases that may dysregulate the immune response could be present, including that of previous SARS-CoV-2 infection, and they are not simultaneously taken into account to determine which has the strongest explanatory power. We were therefore motivated to pursue careful multivariate statistical regression analysis to attempt to distinguish which variables have a statistically stronger influence in driving humoral decay kinetics when many variables are considered simultaneously.

The lasso regression analysis, presented in Fig. 3, highlights a dynamic shift in the significance of these variables. In the context of the booster dose, the influence of senior and resident status decreases earlier on the lasso path, while age persists as a consistent determinant of decay rates. Conversely, the primary series exhibits an inverse trend, where the contribution of age diminishes under increasing penalization, and the roles of senior and resident status remain prominent. This trade-off between age and residency, or senior status, suggests that these factors are not completely distinguishable within our dataset. The strong positive correlation observed between age and senior status, with a correlation coefficient of 0.62, further underscores this complexity. It indicates that the roles of age and status (resident, senior, or HCW) may overlap or that one may serve as a proxy for the other, potentially obscuring other unidentified factors (e.g., the presence of non-communicative diseases) that contribute to the observed patterns. Decoupling the affects of immunosenescence on immune kinetics as a function of age from other potential influences remains challenging. Our multivariate linear regression analysis emphasizes the significance of age in shaping immune responses, even when controlling for other comorbidities. We maintain that chronological age is a strong proxy for more rapid decay kinetics (see fig. 1C and fig. 2A). However, decoupling age-related dysregulation of the innate and adaptive immune system and remodeling of immune organ structure that accumulate with aging, remains a challenge.

In summary, we find that an LNP mRNA-based booster dose leads to a 71-84% increase in Anti-S half life relative to the primary series Anti-S half life. Sex at birth appears to be an interesting feature, whereby our single-variate statistical test shows no significant difference in decay kinetics between males and females (Fig. 2A), despite this result, our multi-variate approach reveals males to have statistically significant, albeit small, bias in favour of increased Anti-S half life (Fig 4). We are able to reproduce the recent observations that individuals with pre-existing asthma have an increased vaccine-elicited humoral half life. Consistent with previous literature we also find individuals with non-asthma-related chronic lung disease to have an impaired humoral response. The Anti-S half life following the primary series and booster dose is revealed to decrease as a function of increased chronological age (Fig 1C), although the affect, statistically significant, is minute.

## Data Availability

Data used for this work was acquired through a COVID-19 Immunity Task Force Grant. Requisitions to access the data must be made through the COVID-19 Immunity Task Force.

## 5 Acknowledgements

CSK acknowledges NSERC postdoctoral funding. The authors thank the COVID-19 Immunity Task Force for consolidating COVID-19 immunological data and facilitating access through public grant opportunities, and specifically thank Dr. Mark Brockman for contributing this data set.

