## Supplementary material for "SARS-CoV-2 booster vaccine dose significantly extends humoral immune response half-life beyond the primary series": Supplmementary material

February 6, 2024

Chapin S. Korosec, David W. Dick, Iain R. Moyles, James Watmough

### S1 Supporting Text

### S2 Observation and error models

Eq. 1 is fit to individual Anti-S trajectories in Monolix. The observation model used in these fits is given by,

*y*_2_ = log(*A*) + (*a* + *b*log(*A*)) ∗ *e,*	(S1)

where *a* and *b* are error model parameters, and *e* is a sequence of independent random variables normally distributed with mean 0 and variance 1. The individual model used to fit the decay rate, *γ_j,i_*, is given by,

log(*γj,i*) = log(*γjpop*) + *ηγj,i,*	(S2)

where *η_γj,i_* are the random effects for the decay rate of the *jth* dose for the *ith* individual.

### S3 Population and individual model fits

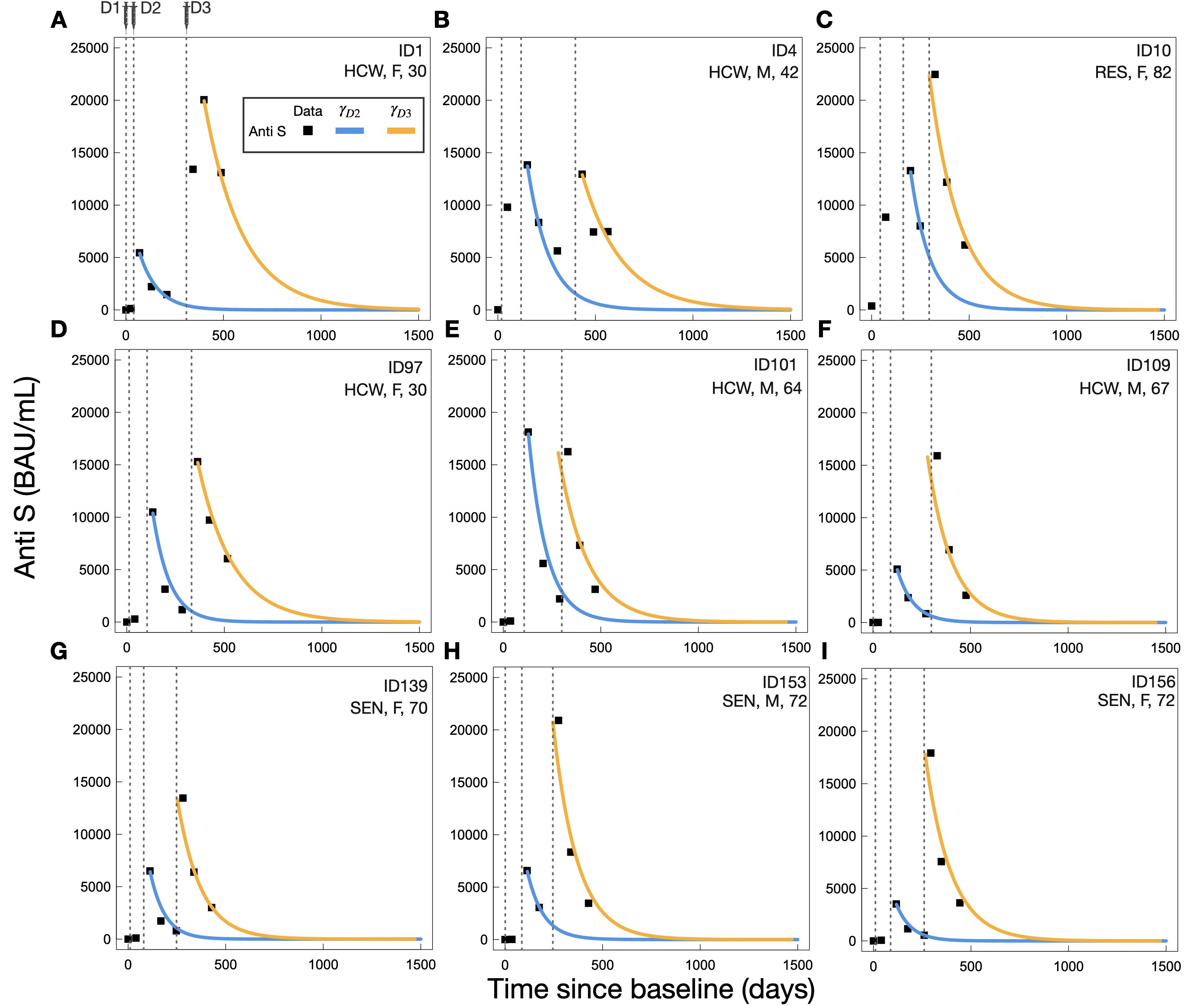

Figure S1: Example individual fits from the primary series and booster dose Anti-S trajectory data. Vertical dashed lines from left to right indicate day of SARS-CoV-2 dose one, two, and three, respectively.

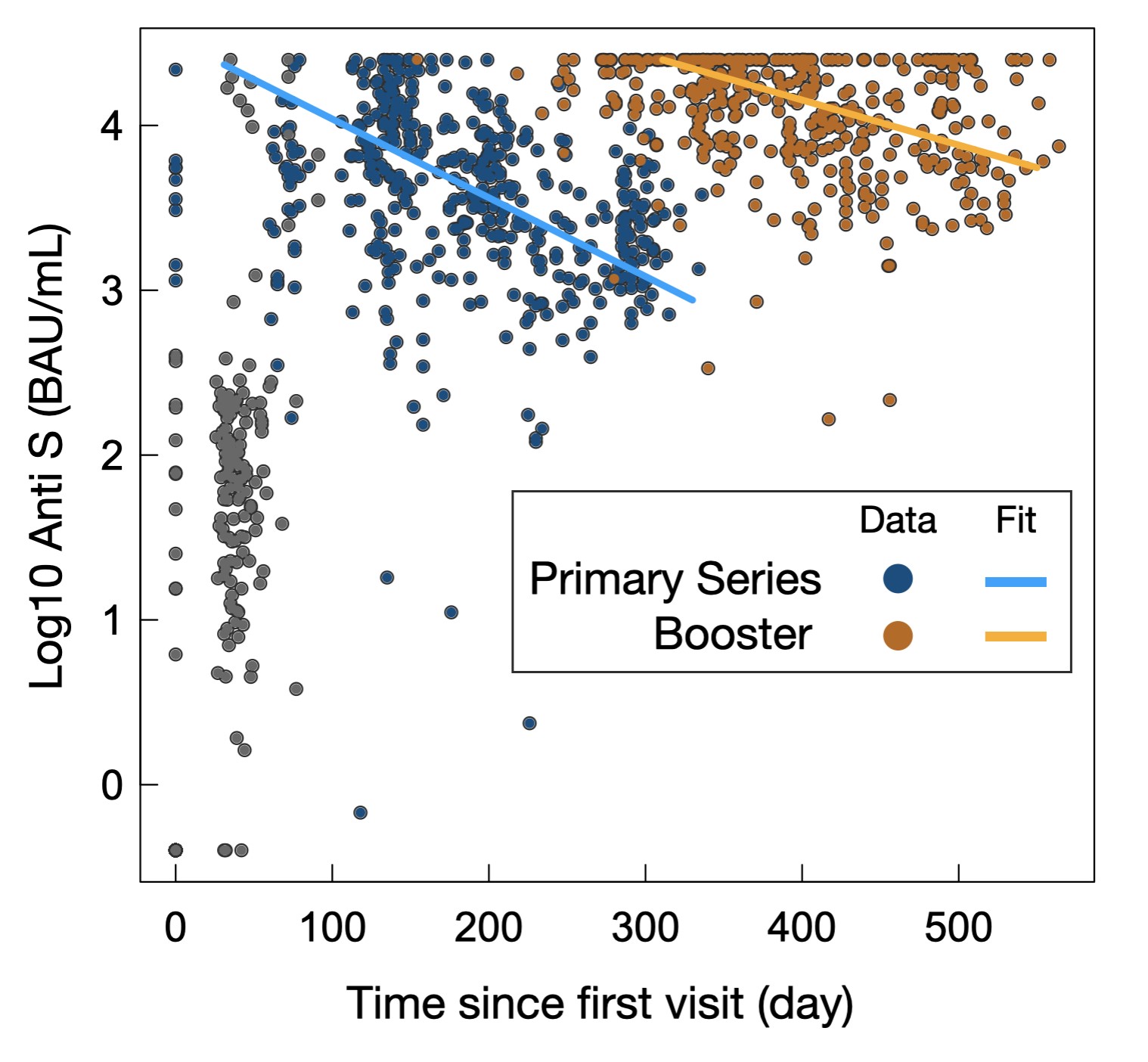

Figure S2: Population fits to all primary series and booster dose data.

### S4 Exploring the age cutoff for statistical comparisons

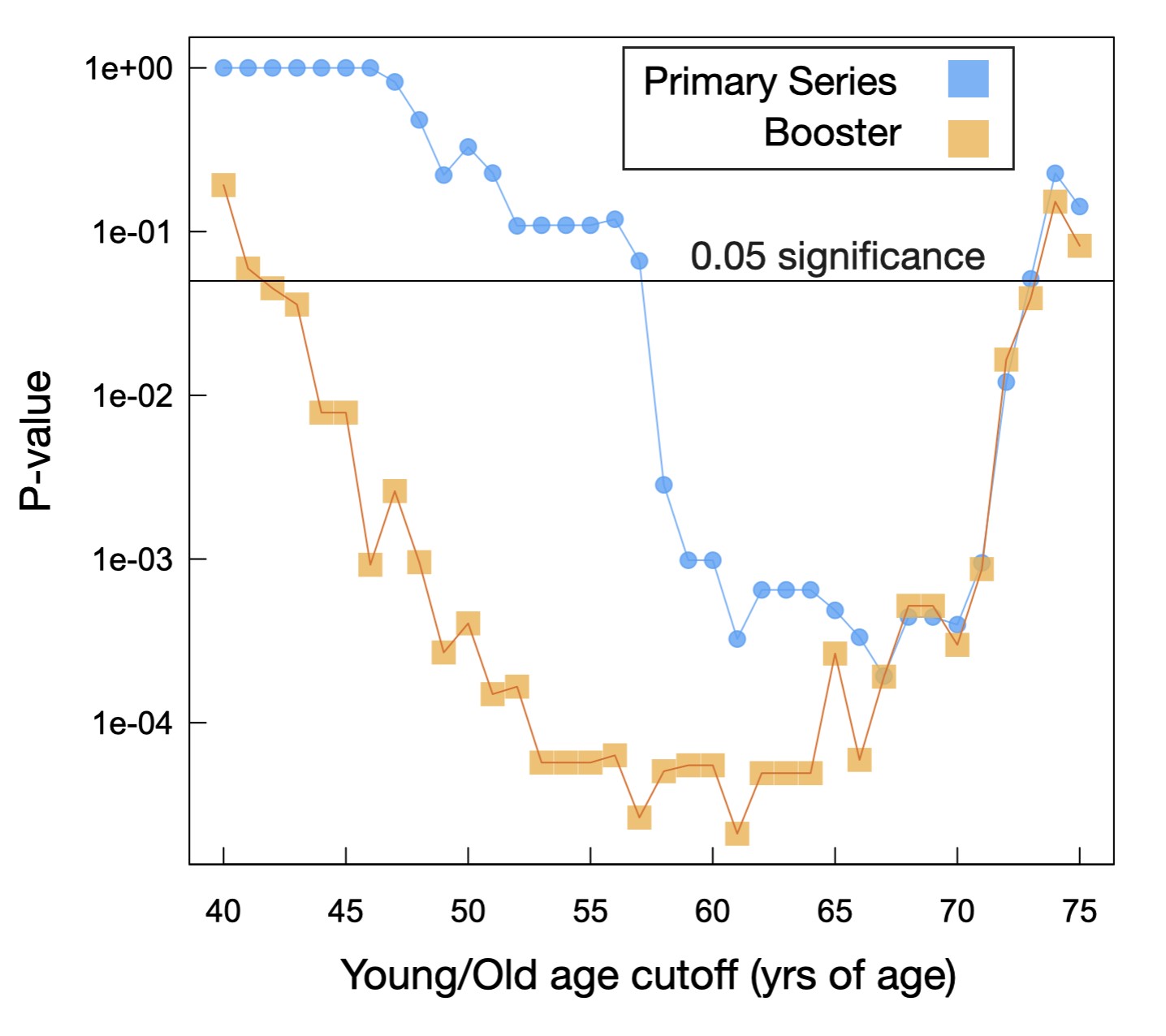

Figure S3: Bonferoni-corrected P-values as a function of young/old age cutoff.

### S5 Multivariate statistical analysis

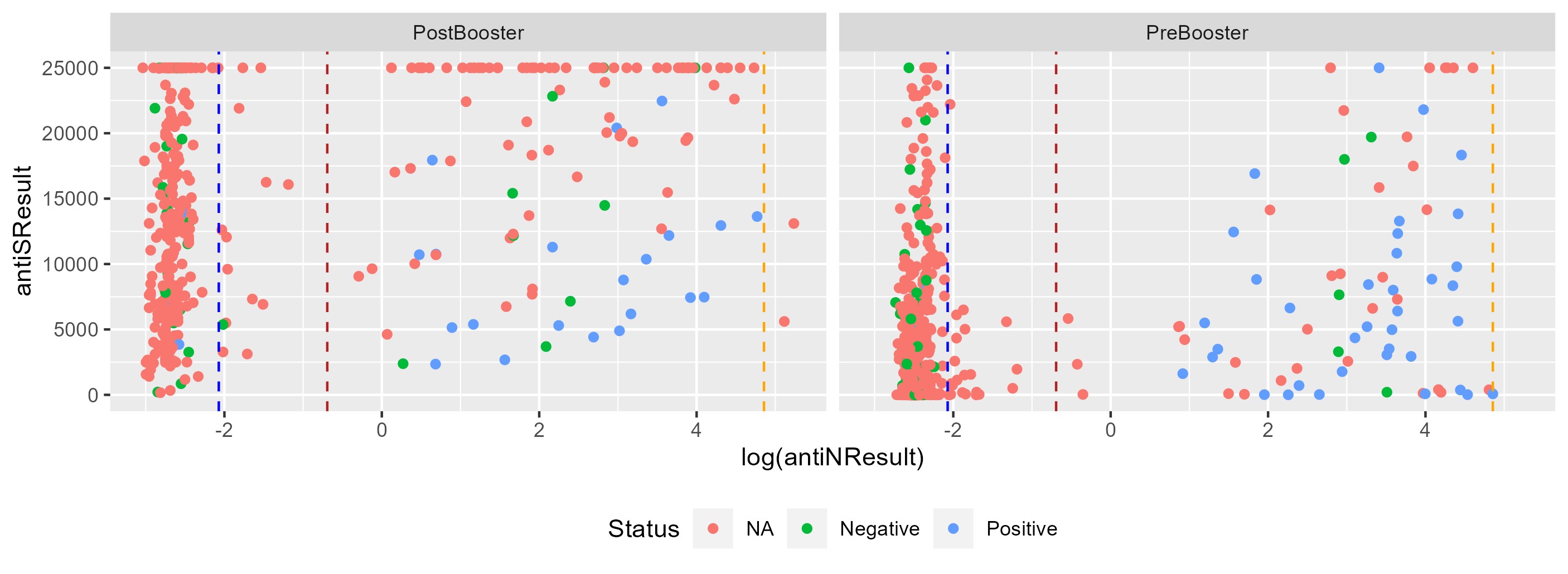

Figure S4: This figure illustrates the relationship between the logarithm of N-terminal domain (anti-N) antibody levels and spike protein (anti-S) antibody levels, with data points colored by COVID-19 test status. Vertical dashed lines represent thresholds used to infer assumed positive status before booster administration: low (dark red), alternative (light red), and high (green). These thresholds are determined by: Alternative Threshold (light red): Represented by the middle vertical dashed line. An arbitrary fixed value, An arbitrary fixed value, capturing the visual nadir in the distribution of anti-N results. Low Threshold (dark red): A value that’s set at twice the lowest N-terminal domain antibody levels observed among those who tested positive for COVID-19. High Threshold (green color): Set by the highest N-terminal domain antibody levels observed among those who have not tested positive for COVID-19.

Panel A and B represent pre- or post-booster dose data points, respectively.

#### S5.1 Lasso Regression

Our MVLR model was developed in R version 4.2.2 (2022-10-31 ucrt) on a x86 64-w64-mingw32/x64 (64-bit) platform, running under Windows 10 x64 (build 22621). For feature selection, Lasso regression was employed using the glmnet 4.1-7 package [1–4].

Lasso (Least Absolute Shrinkage and Selection Operator) regression is a form of linear regression that introduces a penalty term on the absolute values of the regression coefficients *β_j_*. Specifically, the L1 penalty term is given by:

*p*

L1 Penalty = *λ*^X^|*β_j_*|

*j*=1

$$L1 Penalty=\lambda\sum^{p} |\beta_{j}|$$

where *λ* is a non-negative regularization parameter, and *β_j_* represents each coefficient in the linear model.

The L1 penalty encourages sparse solutions by setting some coefficients to zero, effectively performing feature

selection.

| term | estimate | std.error | statistic | p.value | model | conf.low | conf.high | signif.star |
| --- | --- | --- | --- | --- | --- | --- | --- | --- |
| Anti-N Result | -0.000005 | 0.000003 | -1.717728 | 0.086289 | Primary Series | -0.000010 | 0.000001 |  |
| Resident (HCW) | 0.000915 | 0.000149 | 6.135707 | 0.000000 | Primary Series | 0.000623 | 0.001207 | *** |
| Senior (HCW) | 0.000381 | 0.000102 | 3.731346 | 0.000206 | Primary Series | 0.000181 | 0.000580 | *** |
| Sex Male (Female) | -0.000214 | 0.000095 | -2.255650 | 0.024401 | Primary Series | -0.000400 | -0.000028 | * |
| Chronic Neurological Disorder | 0.001029 | 0.000188 | 5.476516 | 0.000000 | Primary Series | 0.000661 | 0.001398 | *** |
| Chronic Lung Disease | 0.001873 | 0.000221 | 8.468033 | 0.000000 | Primary Series | 0.001439 | 0.002306 | *** |
| Cancer | 0.000150 | 0.000235 | 0.636438 | 0.524699 | Primary Series | -0.000311 | 0.000610 |  |
| predicted infection (pre booster) | -0.000419 | 0.000143 | -2.921897 | 0.003591 | Primary Series | -0.000699 | -0.000138 | ** |
| Anti-N Result | 0.000007 | 0.000003 | 2.117883 | 0.034572 | Booster Dose | 0.000001 | 0.000014 | * |
| Anti-Spike Censored | -0.000856 | 0.000184 | -4.652644 | 0.000004 | Booster Dose | -0.001216 | -0.000495 | *** |
| Resident (HCW) | 0.000175 | 0.000247 | 0.707532 | 0.479494 | Booster Dose | -0.000310 | 0.000660 |  |
| Senior (HCW) | 0.000002 | 0.000196 | 0.010361 | 0.991737 | Booster Dose | -0.000382 | 0.000386 |  |
| Sex Male (Female) | -0.000367 | 0.000108 | -3.401200 | 0.000713 | Booster Dose | -0.000579 | -0.000156 | *** |
| Min-Max Normalized Age | 0.000846 | 0.000356 | 2.376604 | 0.017767 | Booster Dose | 0.000148 | 0.001544 | * |
| Hypertension | 0.000325 | 0.000136 | 2.392787 | 0.017010 | Booster Dose | 0.000059 | 0.000591 | * |
| Asthma | -0.000783 | 0.000179 | -4.369616 | 0.000015 | Booster Dose | -0.001134 | -0.000432 | *** |
| Chronic Lung Disease | 0.004135 | 0.000275 | 15.019343 | 0.000000 | Booster Dose | 0.003595 | 0.004675 | *** |
| Cancer | 0.000494 | 0.000255 | 1.938031 | 0.053061 | Booster Dose | -0.000006 | 0.000994 |  |
| predicted infection (any phase) | -0.000583 | 0.000176 | -3.315878 | 0.000965 | Booster Dose | -0.000928 | -0.000239 | *** |

Table S1: Multivariate Linear Regression Coefficients and Significance

In R the regression objective function is:

Minimize:
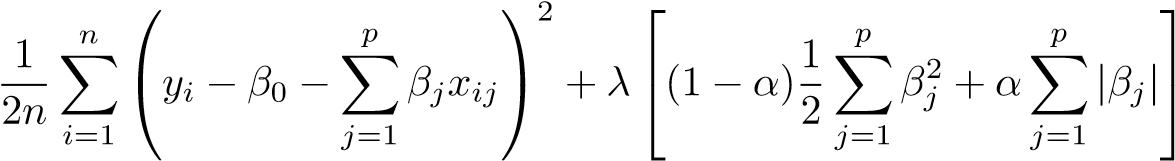

Here: -
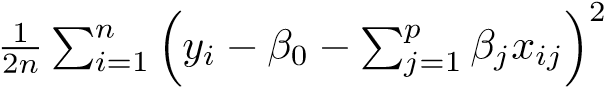
 is the loss term, which measures how well the model fits the data. -

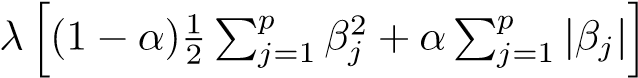
 is the penalty term. *λ* is the regularization parameter, and *α* is the mixing

parameter between L1 and L2 penalties.

For the ’glmnet’ package the parameter *α* controls the balance between Lasso (L1 penalty) and Ridge (L2 penalty). When *α* = 1, it’s Lasso regression. When *α* = 0, it’s Ridge regression. For 0 *< α <* 1, it’s Elastic Net regression, which combines both L1 and L2 penalties. In our case *α* = 1 and we are using Lasso regression. [1]

For *λ* selection the dataset is divided into ’k’ subsets, and the Lasso model is trained on ’k-1’ of these subsets and validated on the remaining one, cycling through all ’k’ subsets and averaging the validation errors to select the best lambda.

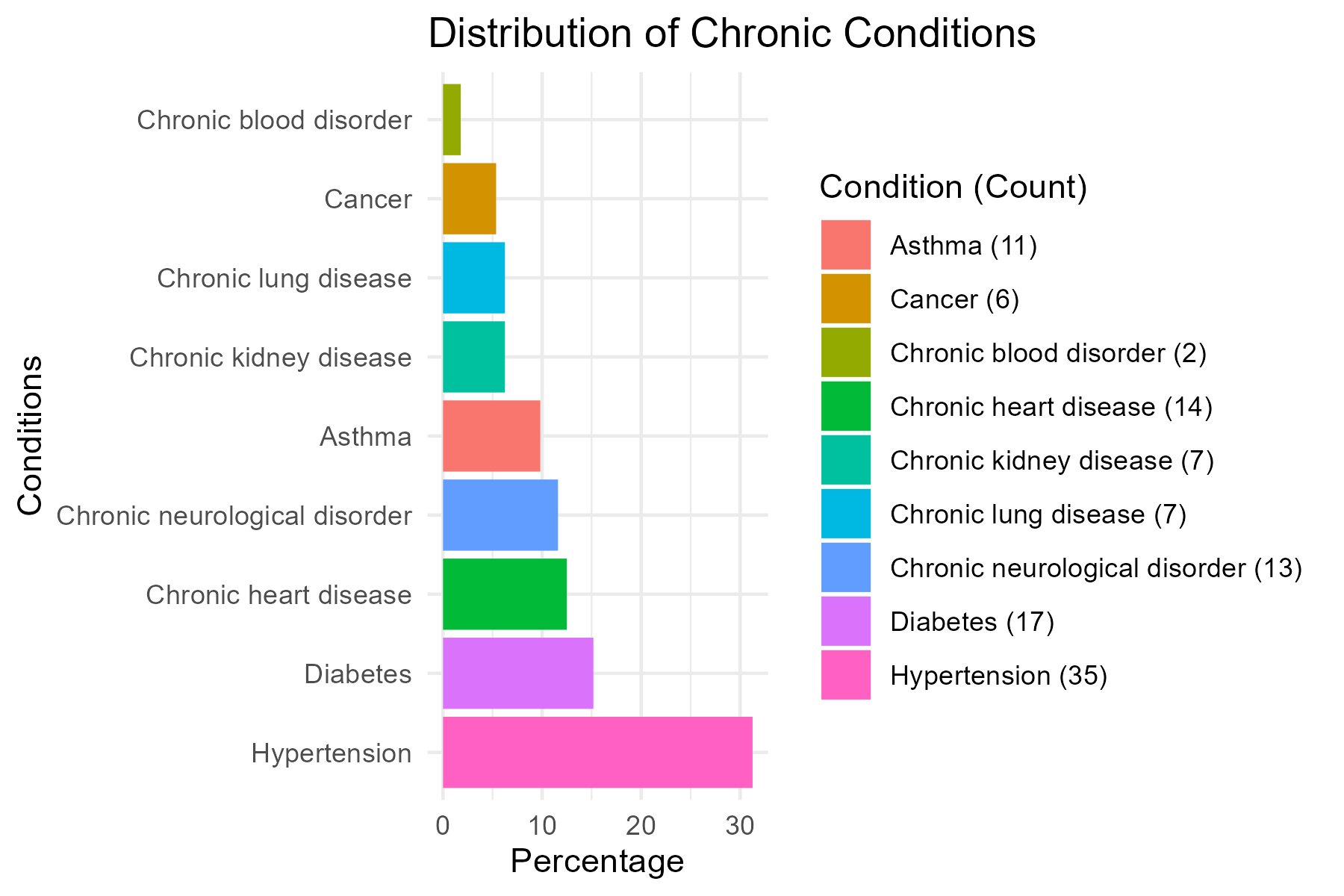

Figure S5: Distribution of chronic conditions among individuals. The bars represent the percentage of diagnoses. The legend details the count of each condition.

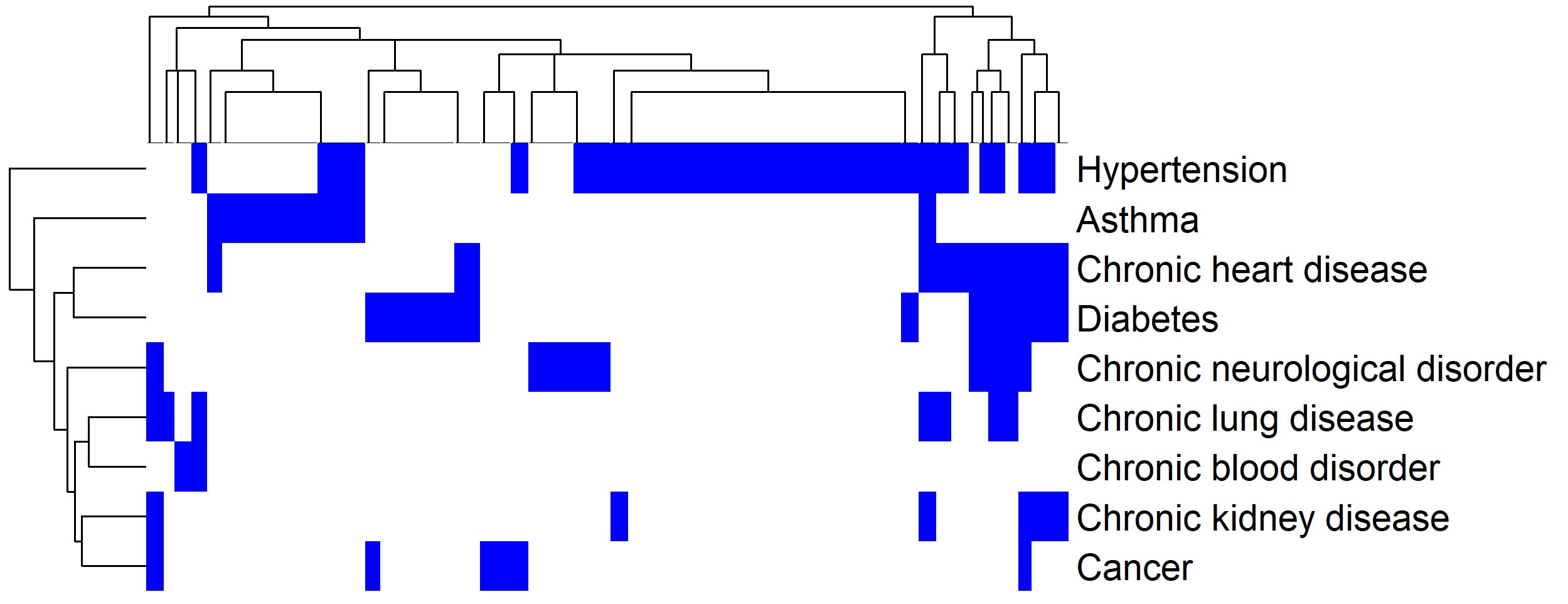

Figure S6: Clustered heatmap illustrating the co-occurrence patterns of chronic comorbidities. Blue cells represent the presence of a chronic comorbidity, with dendrograms showcasing hierarchical clustering based on chronic comorbidity co-occurrence. The counts for each comorbidity are as follows: Hypertension (248), Diabetes (107), Chronic heart disease (88), Asthma (81), Chronic neurological disorder (75), Chronic lung disease (49), Chronic kidney disease

(47), Cancer (43), and Chronic blood disorder (15).

dose2Mu

dose3Mu

normDose2Mu

normDose3Mu

antiNResult

antiSCensor

TypeshortHCW

TypeshortRES

TypeshortSEN

groupTypeNonHCW

SexatBirthMale

assumedPositive

assumedPositiveInd0

assumedPositivePreBoosterInd1

ageMinMaxNormalized

ChronicneuroDisorder1

Hypertension1

Asthma1

ChroniclungDisease1

ChronickidneyDisease1

ChronicheartDisease1

ChronicbloodDisorder1

Cancer1

Diabetes1

dose2Mu

10.7710.77-0.44-0.32-0.570.660.290.57-0.24-0.330.33-0.20.520.770.45-0.170.760.460.660.150.390.6

dose3Mu

0.7710.771-0.57-0.55-0.550.560.320.55-0.24-0.50.5-0.250.550.660.55-0.250.790.410.550.490.440.55

normalizedDose2Mu

10.7710.77-0.44-0.32-0.570.660.290.57-0.24-0.330.33-0.20.520.770.45-0.170.760.460.660.150.390.6

normalizedDose3Mu

0.7710.771-0.57-0.55-0.550.560.320.55-0.24-0.50.5-0.250.550.660.55-0.250.790.410.550.490.440.55

antiNResult

-0.44-0.57-0.44-0.5710.50.44-0.32-0.34-0.440.250.86-0.860.54-0.47-0.42-0.44-0.03-0.26-0.25-0.33-0.23-0.37-0.4

antiSCensor

-0.32-0.55-0.32-0.550.510.24-0.3-0.11-0.2400.58-0.580.11-0.28-0.42-0.360.1-0.31-0.22-0.26-0.2-0.34-0.28

TypeshortHCW

-0.57-0.55-0.57-0.550.440.241-0.61-0.86-10.020.35-0.350.14-0.98-0.58-0.650-0.47-0.48-0.62-0.02-0.53-0.57

TypeshortRESIDENT

0.660.560.660.56-0.32-0.3-0.6110.110.610.03-0.160.160.040.620.840.63-0.130.660.780.83-0.10.390.76

TypeshortSENIOR

0.290.320.290.32-0.34-0.11-0.860.1110.86-0.04-0.330.33-0.190.830.180.410.090.160.10.240.090.410.22

groupTypenon-HCW

0.570.550.570.55-0.44-0.24-10.610.861-0.02-0.350.35-0.140.980.580.6500.470.480.620.020.530.57

SexatBirthMale

-0.24-0.24-0.24-0.240.2500.020.03-0.04-0.0210.26-0.260.290-0.15-0.030.02-0.02-0.020.05-0.24-0.31-0.13

assumedPositivePositive

-0.33-0.5-0.33-0.50.860.580.35-0.16-0.33-0.350.261-10.71-0.37-0.31-0.390.06-0.11-0.03-0.18-0.26-0.29-0.27

assumedPositiveIndicator0

0.330.50.330.5-0.86-0.58-0.350.160.330.35-0.26-11-0.710.370.310.39-0.060.110.030.180.260.290.27

assumedPositivePreBoosterIndicator1

-0.2-0.25-0.2-0.250.540.110.140.04-0.19-0.140.290.71-0.711-0.15-0.1-0.2300.050.13-0.03-0.21-0.13-0.13

ageMinMaxNormalized

0.520.550.520.55-0.47-0.28-0.980.620.830.980-0.370.37-0.1510.560.730.050.460.510.6300.510.57

Chronicneurologicaldisorder1

0.770.660.770.66-0.42-0.42-0.580.840.180.58-0.15-0.310.31-0.10.5610.46-0.230.680.570.71-0.020.490.69

Hypertension1

0.450.550.450.55-0.44-0.36-0.650.630.410.65-0.03-0.390.39-0.230.730.4610.070.470.480.580.120.280.44

Asthma1

-0.17-0.25-0.17-0.25-0.030.10-0.130.0900.020.06-0.0600.05-0.230.071-0.11-0.020-0.16-0.19-0.23

Chroniclungdisease1

0.760.790.760.79-0.26-0.31-0.470.660.160.47-0.02-0.110.110.050.460.680.47-0.1110.540.690.340.290.48

Chronickidneydisease1

0.460.410.460.41-0.25-0.22-0.480.780.10.48-0.02-0.030.030.130.510.570.48-0.020.5410.75-0.140.50.63

Chronicheartdisease1

0.660.550.660.55-0.33-0.26-0.620.830.240.620.05-0.180.18-0.030.630.710.5800.690.751-0.10.30.83

Chronicblooddisorder1

0.150.490.150.49-0.23-0.2-0.02-0.10.090.02-0.24-0.260.26-0.210-0.020.12-0.160.34-0.14-0.11-0.03-0.09

Cancer1

0.390.440.390.44-0.37-0.34-0.530.390.410.53-0.31-0.290.29-0.130.510.490.28-0.190.290.50.3-0.0310.42

Diabetes1

0.60.550.60.55-0.4-0.28-0.570.760.220.57-0.13-0.270.27-0.130.570.690.44-0.230.480.630.83-0.090.421

TableS2:Correlationrelationshipsbetweenvariouschroniccommodities,demographicfactors.

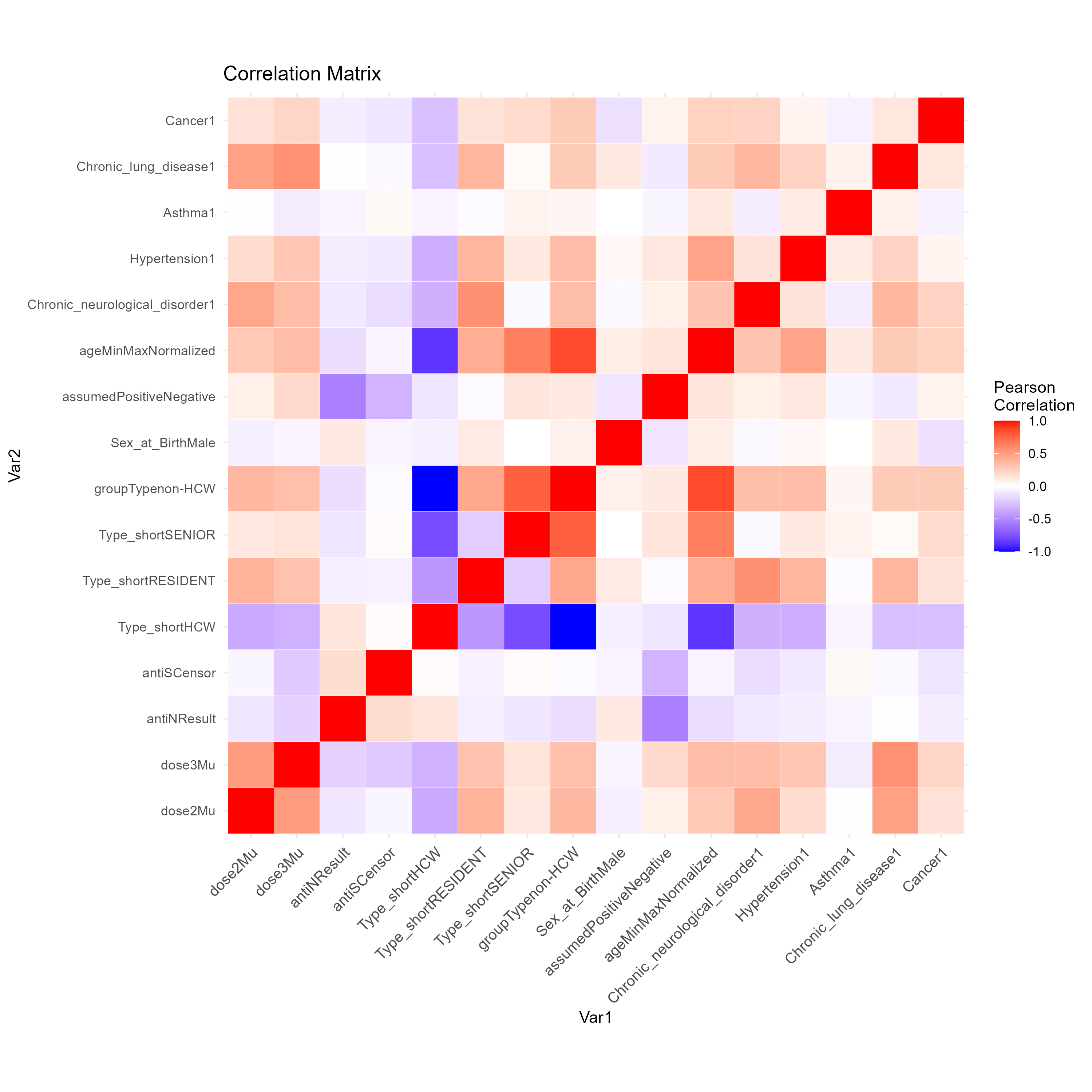

Figure S7: Correlation heatmap illustrating relationships between various chronic commodities, demographic factors, and vaccine dose responses available in the data. Red indicates positive correlation, blue indicates negative correlation, and white indicates no correlation.

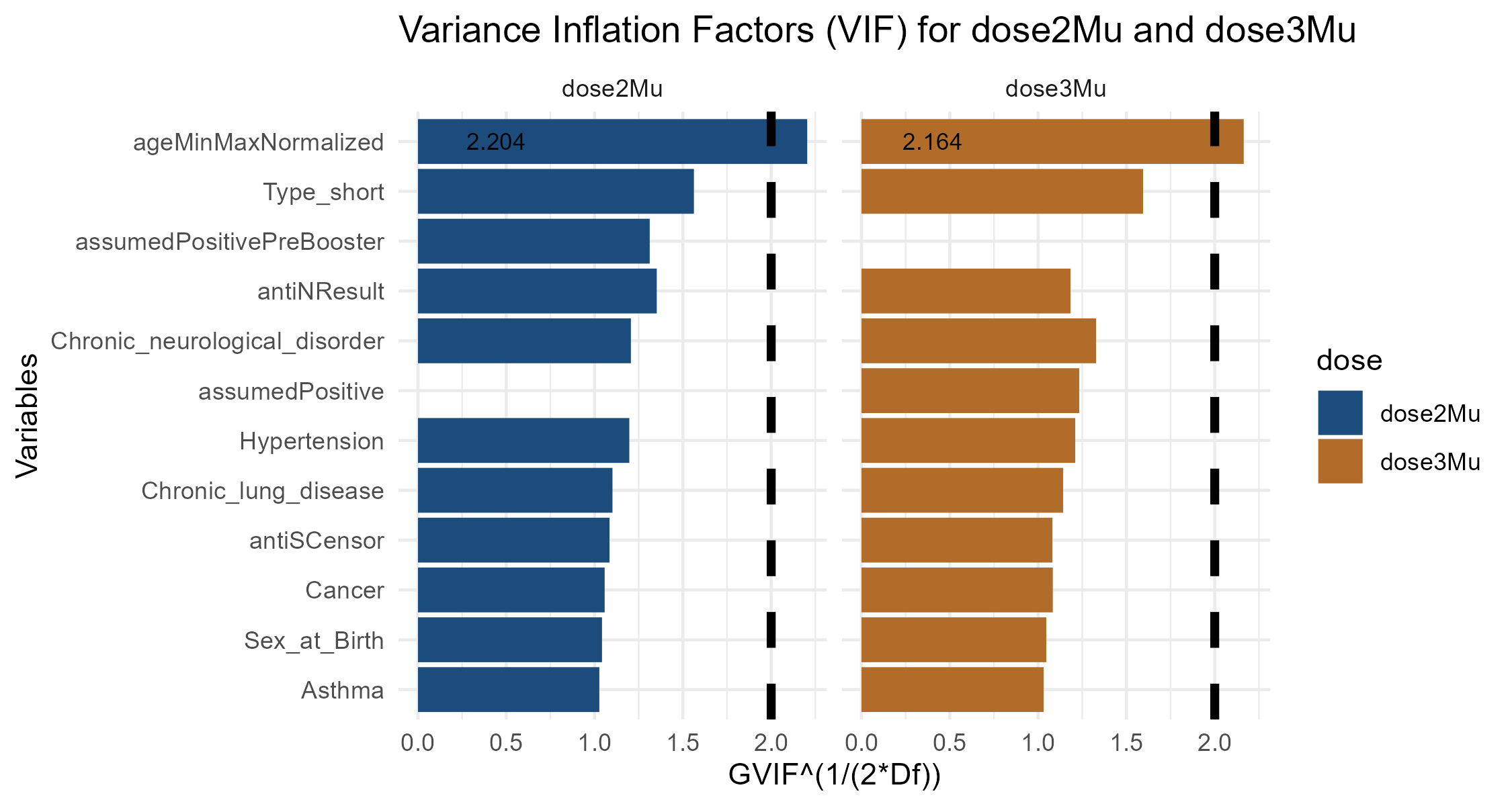

Figure S8: Variance Inflation Factor (VIF) Analysis. The figure displays GVIF^(1^*^/^*^(2∗^*^Df^*^))^ values for each variable considerd in the regression models. All variables, except for ‘ageMinMaxNormalized‘, exhibit values below the threshold of 2. This suggests that there are no substantial multicollinearity concerns within these models, with the exception of the ‘ageMinMaxNormalized‘ variable. In the VIF analysis, the variable ‘ageMinMaxNormalized‘ exhibited a marginally high GVIF value (2.20 for ‘dose2Mu‘ and 2.16 for ‘dose3Mu‘), just slightly above the conventional threshold of 2 [5–7]. This prompts a discussion on whether to include or exclude this variable from the model. The slight exceedance of the threshold does not necessarily denote a substantial violation of the assumptions underlying the model. The variable ‘ageMinMaxNormalized‘ is the focus of our analysis and excluding it based solely on a marginally high VIF value could lead to an incomplete or biased understanding of the underlying relationships. For this reason we include ‘ageMinMaxNormalized‘ in the analysis.

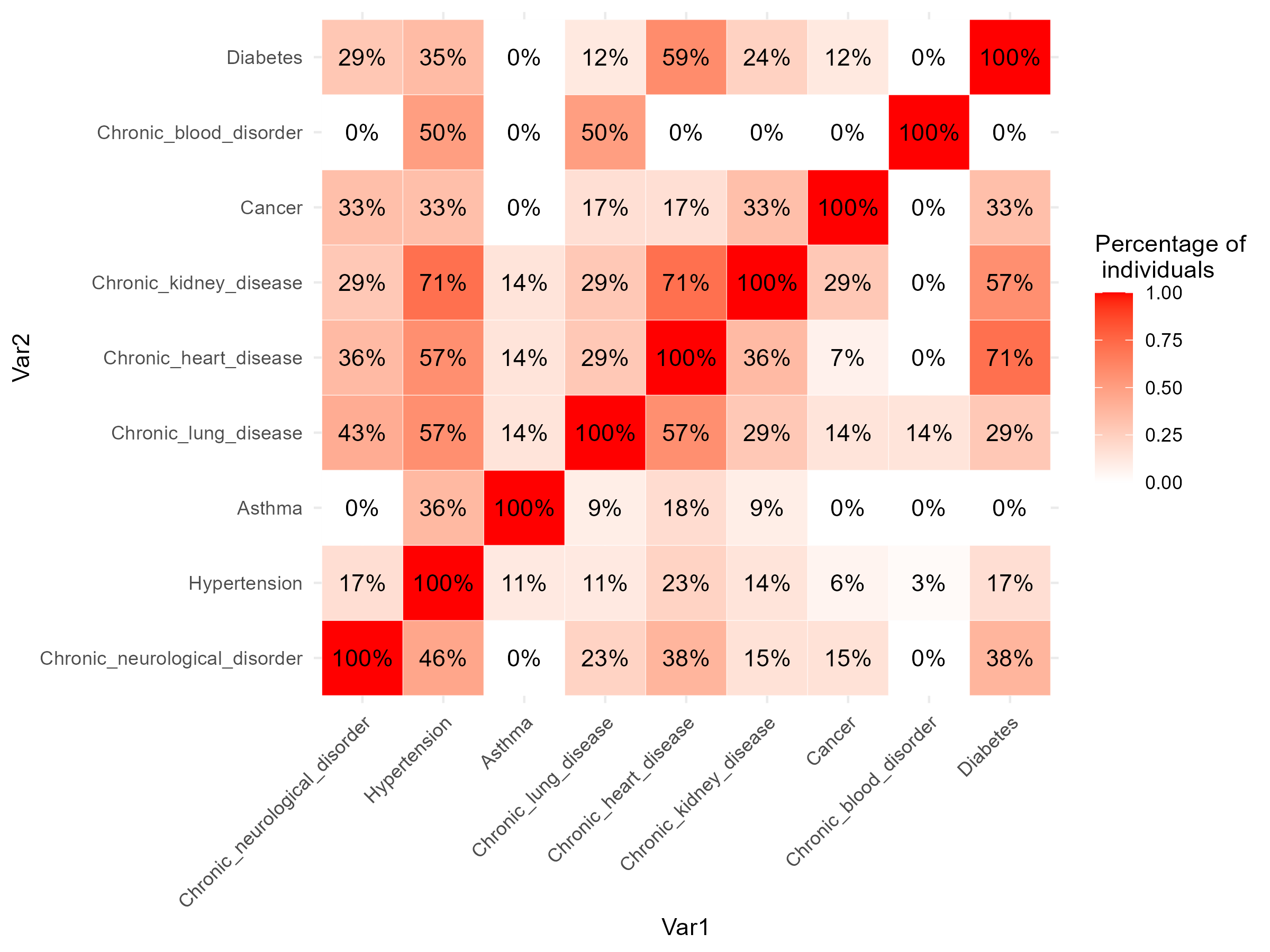

Figure S9: The heatmap portrays the percentage of individuals in our dataset with co-existing chronic diseases. Each cell represents the proportion of individuals having a specific condition (represented by columns) given the presence of another condition (indicated by rows). The color gradient corresponds with the percentage: deeper red shades signify a higher percentage, while white denotes a lower one. This represents the conditional probability, in percentage terms, that an individual possesses the comorbidity listed in the row, given they have the one in the column. For example, if someone has Hypertension (row), the probability they also have Hypertension (column) is 100%. However, the probability that they have Diabetes (column) drops to 17%. The repetition of percentages across different conditions can be attributed to the limited count of individuals with certain diseases in the dataset.
